# Machine learning approach to assess the pathogenicity of BRCA1/2 genetic variants : brca-NOVUS

**DOI:** 10.1101/2023.10.20.23297295

**Authors:** Aastha Vatsyayan, Vinod Scaria

## Abstract

Breast cancer is globally the leading type of cancer in terms of both incidence and mortality. BRCA1 and BRCA2 gene variants have long been linked to and studied in context of the disease. Rapid variant discovery has further been made freely accessible by advances in Next-generation sequencing, making it a demanding task to accurately interpret these variants for clinical and research applications. To establish the nature of these variants, the American College of Medical Genetics and Genomics and the Association of Molecular Pathologists (ACMG-AMP) have issued a set of guidelines for variant classification. However, given the huge number of variants associated with the two large and well-studied genes, functional studies or ACMG-AMP classification is a mountainous challenge. Here we describe brca-NOVUS, a machine learning approach trained on a gold-standard ACMG-qualified dataset for the accurate interpretation of variants at large scale. Using two independent test and validation datasets of ACMG-qualified variants, we show that brca-NOVUS can be used to for the classification of variants in clinical as well as research settings.

## INTRODUCTION

Female Breast cancer is the most commonly diagnosed cancer globally, and is the leading cause of cancer death in women, according to the GLOBOCAN 2020^1^ data. Early diagnosis can greatly increase survival outcomes, however regular screening may neither be indicated, nor be possible for all. The presence of causative variants in the BRCA genes puts a carrier at a higher lifetime risk of cancer than the general population - up to 72% of women with pathogenic mutations will develop breast cancer by 70–80 years of age^2^. Thus population screening can lead to the identification of at-risk individuals who can then go in for regular screening procedures. The accurate interpretation of genetic variants plays a crucial role here in determining the course of action for potential carriers.

Since the discovery of BRCA1 and BRCA2 about 30 years ago^3^, significant efforts have gone into studying the genes and their variants causing breast and ovarian cancer. Advances in Next-generation sequencing (NGS) technology and its increasing affordability have enabled newer variants to be detected and discovered at a very fast pace. However, the accurate classification of these variants presents a huge challenge for functional approaches to tackle at a commensurate rapid pace. While the American College of Medical Genetics and Genomics and the Association of Molecular Pathologists (ACMG-AMP) have issued a set of guidelines for accurate variant classification, their implementation is a time intensive and expertise-dependant process.

Machine learning approaches in clinical medicine have greatly increased our ability to analyse and interpret large-scale medical data in a number of scenarios. Here we describe a machine learning model trained on ACMG-AMP qualified data that can be used to rapidly classify large numbers of variants. We validate the model on an independent ACMG-qualified validation set. We further demonstrate the model’s utility by classifying and comparing an independent dataset of variants classified by an expert panel following the ENIGMA classification guidelines with the predictions generated by our tool. A public implementation of the algorithm is available at https://github.com/aastha-v/brca-NOVUS.

## MATERIALS AND METHODS

### Datasets

The ClinVar^4^ database was queried for BRCA1 and BRCA2 variants that had been annotated as per the ACMG/AMP guidelines.

### Training Dataset

Variants that were definitively classified as either Pathogenic or Benign by the ClinVar expert panel according to the ACMG & AMP guidelines were collected for model training. The datasets contained a total of 3340 BRCA1 and 4107 BRCA2 variants based on the human genome hg38 assembly. All non-exonic variants, Variants of Uncertain Significance, and variants that were not SNVs were removed. This resulted in a total of 2753 BRCA1 variants, of which 2189 were annotated as Pathogenic / Likely Pathogenic, and 563 as Benign / Likely Benign. Similarly for BRCA2, out of 3492 remaining variants, 2629 were annotated as Pathogenic / Likely Pathogenic, and 863 as Benign / Likely Benign.

### Test Dataset

The remaining variants from the ClinVar dataset that had been ACMG/AMP annotated, but had not yet been reviewed by the expert panel were collected for the Test dataset. Out of 8802 BRCA1 variants, 1577 exonic SNVs that were not Variants of Uncertain Significance were collected, out of which 945 were Pathogenic / Likely Pathogenic and 632 were Benign / Likely Benign. For BRCA2, out of 9895 BRCA2 variants, 3492 reminded after processing, out of which 2629 were annotated as Pathogenic / Likely Pathogenic, and 863 as Benign / Likely Benign.

### Independent ACMG-Qualified Validation Dataset

Our validation dataset consisted of variants collected from the IndiGen dataset, that were manually classified as per the ACMG/AMP guidelines. Upon processing, 99 BRCA1 variants (64 Pathogenic/Likely Pathogenic and 35 Benign / Likely Benign), and 91 BRCA2 (37 Pathogenic/Likely Pathogenic and 54 Benign / Likely Benign) were obtained.

### Machine Learning Algorithms

We compared several models from different algorithms for performance, including TabNet^5^, a novel deep learning neural network designed specifically for tabular data, and XGBoost^6^, a scalable tree boosting system. However, tree ensemble models (such as Random Forest) are traditionally the gold standard in classifying tabular data. This was supported by our comparison of model performance, and XGBoost which consistently outperformed TabNet was taken as the algorithm of choice for classifying our variants.

### Setup and Data Preprocessing

For preprocessing, local installations of ANNOVAR^7^ and Ensembl’s Variant Effect Predictor^8^ (VEP) tools and data from the UCSC Genome Browser database^9^ were utilised. For model running, Anaconda^10^ was used to enable the use of Scikit-learn^11^, Pandas^12^, Matplotlib^13^ and Seaborn^14^ for analysis and visualisation.

### Computing Parameters

A total of 73 attributes were used for model building. These are summarised in **Supplementary Table 1**. A number of genomic parameters were computed using the ANNOVAR software, including allele frequencies from global datasets (GnomAD^15^, 1000Genomes^16^ and GME^17^), along with pathogenicity scores from several tools (SIFT^18^, CADD^19^ etc). Further parameters were computed and encoded using bespoke scripts, including positions of both the nucleotide and protein changes, information if the variant was a high confidence Loss of Function variant in the canonical transcript, information about whether the variant fell into a Pfam important domain, as well as what its exonic function was (e.g. Stopgain/Startloss, frameshift insertion/deletion etc.). Finally, all Pathogenic/Likely Pathogenic variants were encoded as “1”, and all Benign/Likely Benign variants as “0”. The scripts are available on github.

**Table 1:** Comparison between brca-NOVUS and BRCA-ML using different metrics.

|  | BRCA1 |  | BRCA2 |  |
| --- | --- | --- | --- | --- |
|  | brca-NOVUS | BRCA-ML | brca-NOVUS | BRCA-ML |
| **Accuracy** | 0.958 | 0.716 | 0.951 | 0.402 |
| **PPV** | 0.986 | 0.989 | 0.947 | 0.925 |
| **NPV** | 0.911 | 0.549 | 0.961 | 0.339 |
| **MCC** | 0.911 | 0.549 | 0.885 | 0.177 |

### Cross Validation

Model hyper-parameters were selected and evaluated using a 5-fold cross validation approach. The data was tested on a number of set splits, including 70% train and 30% test, and 80% train and 20% test sets to enable testing.

### Accuracy Estimates

The following accuracy estimates were used for evaluating the models. a) Sensitivity, b) Specificity c) Accuracy and d) Matthews Correlation Coefficient (MCC).

### Independent Validation Dataset

We queried all BRCA variants from BRCA Exchange, one the the largest publicly accessible repositories of BRCA variants, that offers variant classification by an expert panel based on the ENIGMA classification guidelines. We ran our models on the data, and compared our predictions with the classifications made by the expert panel.

## RESULTS

### ML Models

#### BRCA1

Our best performing BRCA1 model trained on a train-test split of 80-20, and using the following parameters: **scale_pos_weight=3**.**888099467, base_score=0**.**5, booster=‘gbtree’, callbacks=None**,

**colsample_bylevel=1, colsample_bynode=1, colsample_bytree=0**.**3**,

**early_stopping_rounds=None, enable_categorical=False**,

**eval_metric=None, gamma=0**.**4, gpu_id=-1, grow_policy=‘depthwise’**,

**importance_type=None, interaction_constraints=‘‘**,

**learning_rate=0**.**1, max_bin=256, max_cat_to_onehot=4**,

**max_delta_step=0, max_depth=4, max_leaves=0, min_child_weight=1**,

**monotone_constraints=‘()’, n_estimators=150**,

**n_jobs=0, num_parallel_tree=1, predictor=‘auto’, random_state=0**,

**reg_alpha=0, reg_lambda=1, missing=nan, seed=123**

For our training data, our model yielded an accuracy of 0.998, with an AUC of 0.999 and MCC of 0.994. On our test dataset of ClinVar variants, it further gave an accuracy of 98.92%, and an accuracy of 98% on our independent validation IndiGen dataset.

#### BRCA2

For BRCA2, our best model trained on a train-test split of 70-30, using the following parameters: **scale_pos_weight=3**.**046349942, base_score=0**.**5, booster=‘gbtree’, callbacks=None**,

**colsample_bylevel=1, colsample_bynode=1, colsample_bytree=0**.**3**,

**early_stopping_rounds=None, enable_categorical=False**,

**eval_metric=None, gamma=0**.**3, gpu_id=-1, grow_policy=‘depthwise’**,

**importance_type=None, interaction_constraints=‘‘**,

**learning_rate=0**.**2, max_bin=256, max_cat_to_onehot=4**,

**max_delta_step=0, max_depth=15, max_leaves=0, min_child_weight=5**,

**monotone_constraints=‘()’, n_estimators=50, n_jobs=0**,

**num_parallel_tree=1, predictor=‘auto’, random_state=0**,

**reg_alpha=0, reg_lambda=1, missing=nan, seed=124**

It yielded an accuracy of 0.999 on the training data, with an AUC and MCC of 1 and 0.997 respectively. On our ClinVar test data, it classified at an accuracy of 98.36%, and an accuracy of 97.80% on our independent validation IndiGen dataset.

The feature importance, ROC, and Confusion Matrix of each model are shown in **Figures 1,2 and 3**.

**Figure 1:**
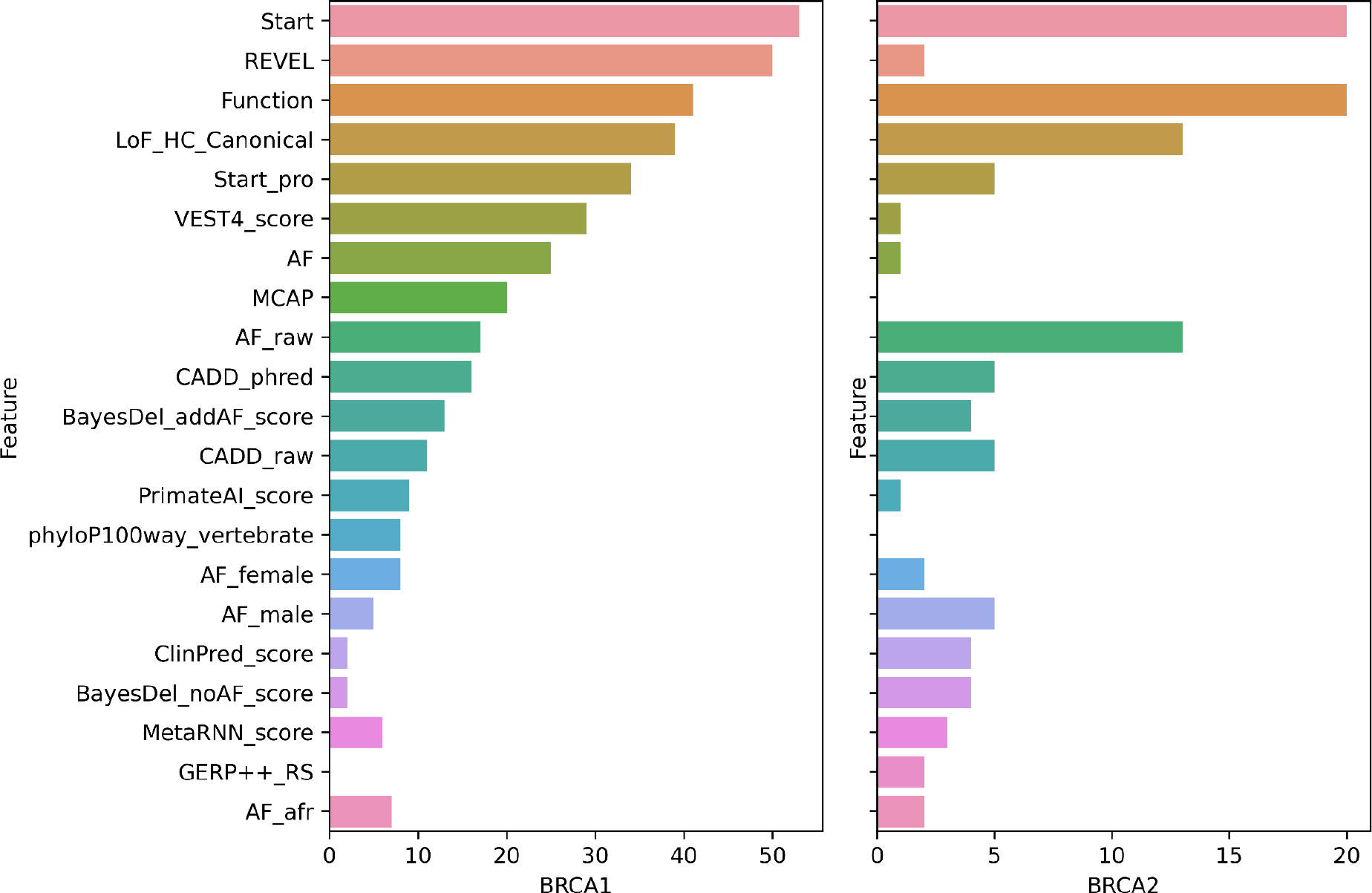
Plot depicting the feature importance of the top 15 features of each gene’s model. The x-axis depicts the F-score for each feature.

**Figure 2:**
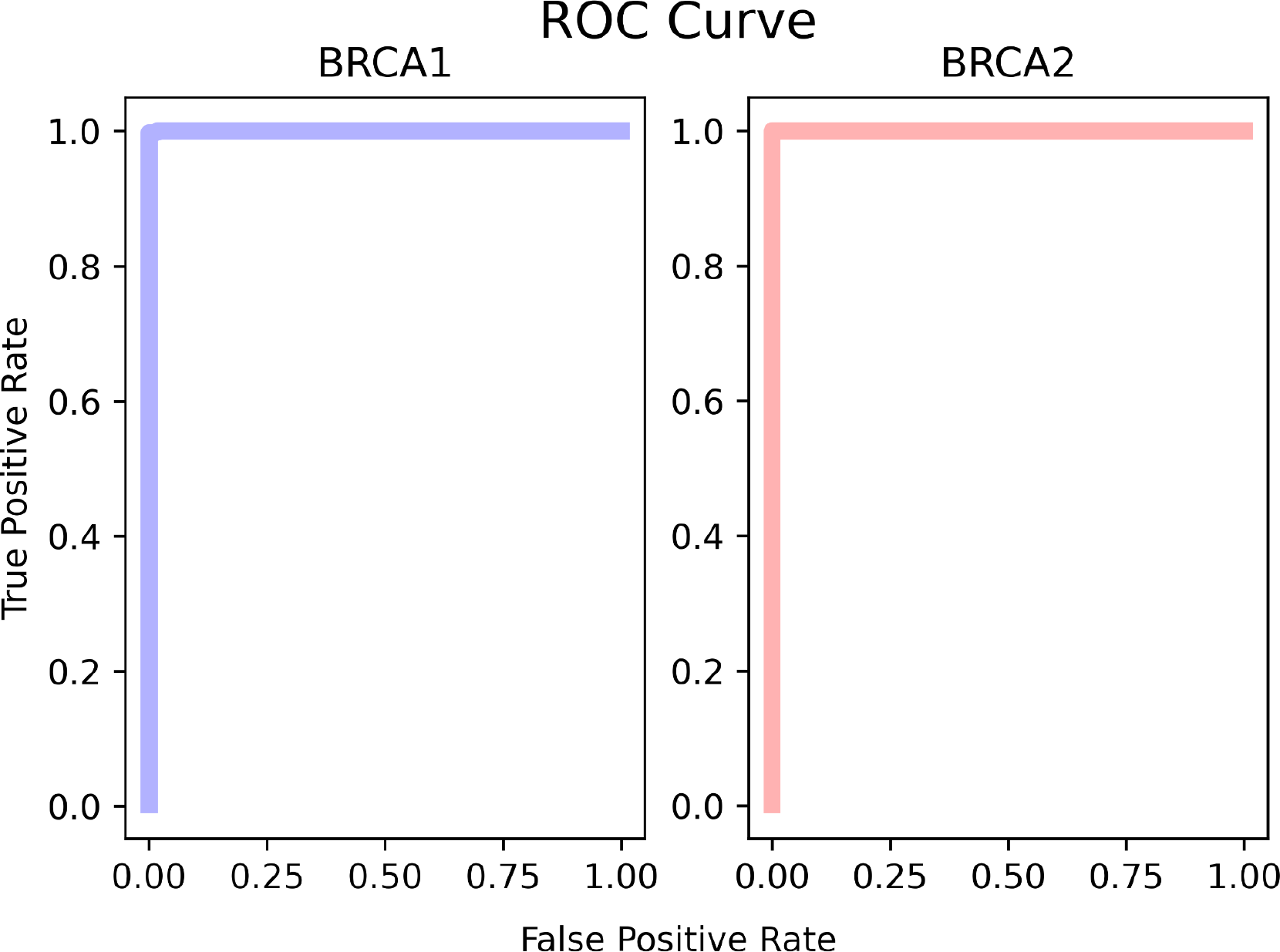
Plot depicting the ROC metric for each gene’s model.

**Figure 3:**
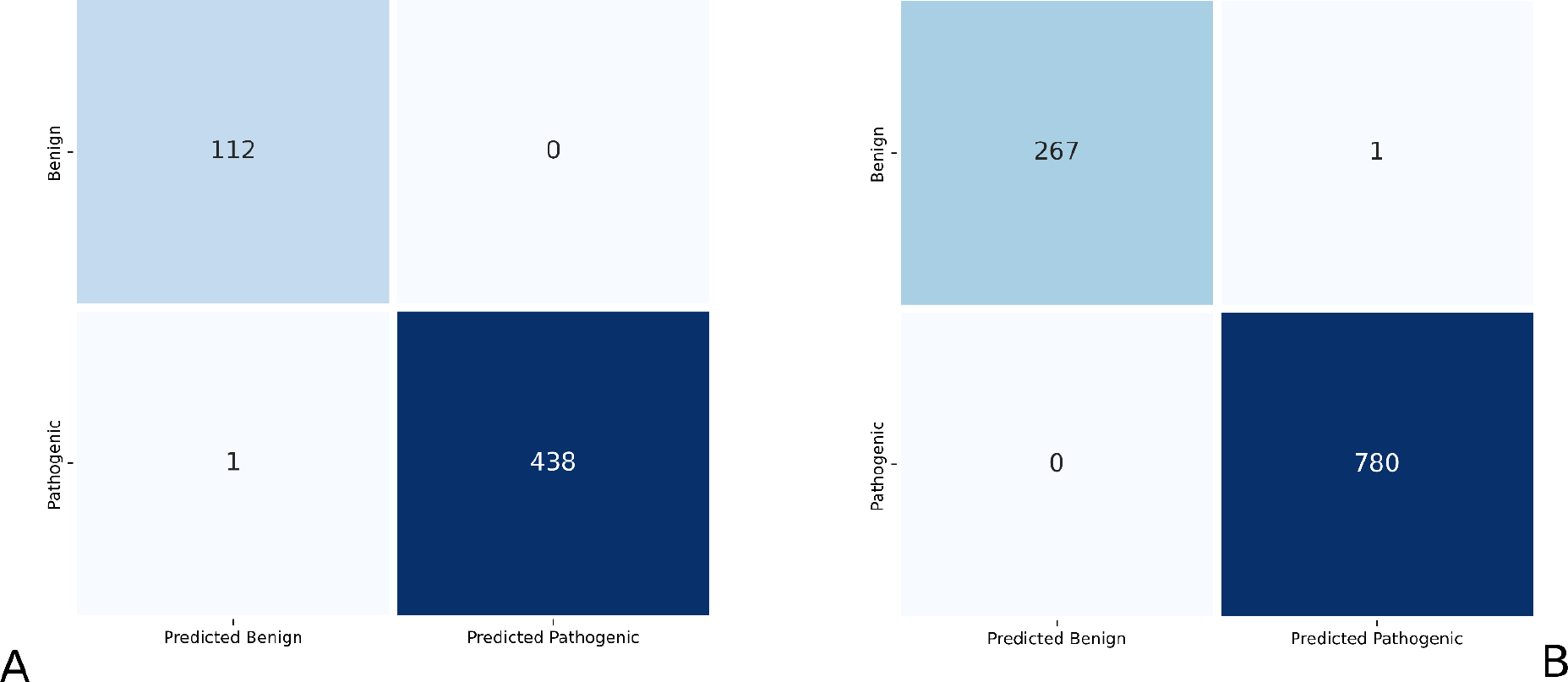
Plot depicting the confusion matrices for (A) BRCA1 and (B) BRCA2 models.

### Comparison with Other Models

To the best of our knowledge, our models are the only models trained on a gold standard dataset of ACMG-AMP qualified BRCA1 and BRCA2 variants. To establish the robustness of the model therefore, we looked at the BRCA-ML^20^ which is trained on data made available by high-throughput functional studies. We compared all variants that were present in our Test dataset as well as had prediction scores generated by their model. We found 613 such variants, 243 belonging to BRCA1 and 370 belonging to the BRCA2 gene. Using the ClinVar classification of these variants as measure, we compared the accuracy of prediction, the positive and negative predictive values (PPV and NPV respectively), and the Matthews Correlation Coefficient (MCC) metrics for each model across both genes. The results are shown in **Table 1**.

The brca-NOVUS models for each gene significantly outperform the corresponding BRCA-ML model across parameters.

### Independent Validation Dataset

We obtained a total of 71,804 BRCA variants from BRCA Exchange, from which we collected 7,445 variants that were ENIGMA expert-classified. We ran our models on each gene, and obtained predictions for 6330 exonic variants. Of 2787 BRCA1 variants, our predictions matched the expert classification for 2786 variants (Accuracy: 99.96412), and of 3543 BRCA2 variants classified, our model’s predictions matched for 3540 variants (Accuracy: 99.91533). The breakdown of the predictions made are given in Table 2. Thus, our models exhibit a high level of accuracy across independently expert classified sets as well.

**Table 2:** Table depicting the number of ENIGMA expert-classified BRCA Exchange variants classified as pathogenic and benign by each of our models.

|  | BRCA1 | BRCA2 |
| --- | --- | --- |
| Benign | 597 | 909 |
| Pathogenic | 2190 | 2634 |
| Total | 2787 | 3543 |

## DISCUSSION

In our work we have shown how brca-NOVUS can be used as an effective tool to classify genetic variants in the BRCA1 and BRCA2 genes. Genetic testing is an essential tool that can be leveraged to successfully identify pathogenic mutation carriers, who can then, through increased surveillance either catch the cancer early and thus increase their chances of survival, or in certain cases prevent the cancer entirely through indicated prophylactic measures. Genetic testing is additionally indicated for family screening, especially with a known family history of cancer. However, genetic testing is only beneficial if an accurate clinical interpretation of the variants can be made. Our work can be used to successfully classify variants of uncertain significance or conflicting interpretations. We have shown the accuracy of our model on an independent ACMG-qualified dataset, as well as an independent ENIGMA-expert panel classified dataset. Additionally, we have also proved the accuracy of both models using a model trained on functional validation studies. The key limitation of our work is that it can only be used to classify exonic variants in both genes, leaving out other variants that may also be disease causing in nature.

## Data Availability

The source code of both our models is available at https://github.com/aastha-v/brca-NOVUS. The models have been standardised on Ubuntu 18 LTS. The instructions and code for the preprocessing pipeline is also included.

## ACKNOWLEDGMENTS

Authors acknowledge funding from the Council of Scientific and Industrial Research (CSIR) through CNP-007 project. The funders had no role in the preparation of the manuscript or decision to publish.

## AUTHOR CONTRIBUTIONS

VS conceptualised, designed and supervised the AI study and algorithm implementation. AV created the code for the implementation of the algorithms, classified the variants, and complied the manuscript.

